# Developing an Early Childhood Environmental Health Vulnerability Index to Assess Cumulative Health Impacts Across Contiguous U.S.

**DOI:** 10.64898/2026.03.10.26348087

**Authors:** Siyu Liu, Anni Yang, Diane Horm, Mingze Zhu, Changjie Cai

## Abstract

Young children (from birth to 5 years old) are uniquely vulnerable to environmental hazards due to their higher exposure relative to body weight, rapid physiological and neurological development, and strong reliance on caregivers for protection and care. Such risks are often amplified in marginalized communities with socioeconomic disadvantage and limited access to resources. However, widely used indices, such as the Social Vulnerability Index (SVI), the Climate Vulnerability Index (CVI) and the Child Opportunity Index (COI), were not specifically developed for young children and may not capture the combined environmental and socioeconomic risks faced by this age group. To address this critical gap, we developed a county-level Early Childhood Environmental Health Vulnerability Index (EC-EHVI) for the contiguous U.S. using multidimensional indicators within an Exposure-Sensitivity-Adaptive Capacity framework and informed by Bronfenbrenner’s bioecological model. We identified the underlying drivers and the spatial patterns of the EC-EHVI. Our results showed that the EC-EHVI exhibited the strongest association with county-level young child mortality and explained a larger proportion of spatial heterogeneity compared with the SVI, CVI, and COI. Elevated vulnerability clustered in the Great Plains and Southeastern U.S., where over half of high-risk counties were exposure-driven, and 411 high–high hotspots were identified. The EC-EHVI offers a valuable spatial decision-support tool for designing targeted, place-based interventions and advancing environmental health equity for young children.

**Plain Language Summary:** Young children (birth to age five) are uniquely vulnerable to environmental hazards. Because their bodies are developing and they consume more air, food, and water relative to their weight, environmental exposures can have severe, lifelong impacts. These risks are often magnified in under-resourced communities. Yet, most existing vulnerability tools were not built with young children in mind, potentially obscuring the combined environmental and social threats they face. To address this gap, we developed a new county-level index to pinpoint where young children are most at risk across the contiguous United States. Our tool integrates data on environmental exposure, community sensitivity, and the resources available to help families cope. When tested, our new index was more strongly linked to young child mortality than several widely used existing measures. We identified major high-risk clusters, particularly in the Great Plains and the Southeastern U.S. This tool can help policymakers and public health officials better target resources and interventions to protect young children and promote environmental health equity.

**Key Points:**

- We developed a county-level Early Childhood Environmental Health Vulnerability Index across the contiguous U.S.
- Elevated vulnerability clustered in the Southeast, Great Plains, and Appalachia, with additional hotspots in Michigan and Maine.
- More than half of high-vulnerability counties were exposure-driven, emphasizing the key role of environmental hazards in child health.

## 1 Introduction

The impact of environmental exposure on human health is increasingly recognized, with extreme climate events and environmental pollution disproportionately affecting marginalized and low-income populations. For example, environmental pollution alone accounts for an estimated 9 million premature deaths annually, which is one in every six worldwide (Landrigan et al., 2018). This burden is not distributed homogeneously among population. Over 92% of pollution-related mortality occurs in low- and middle-income countries (LMICs), highlighting the global disparity in environmental health (Zajac et al., 2025).

Children are highly vulnerable to environmental stressors, with an estimate of more than one in four deaths in children under five attributed to the modifiable environmental risks (World Health Organization (WHO), 2017). From birth to five, children face a period of unique biological vulnerability, as their higher relative intake of air, water, and food per unit of body weight can lead to greater proportional exposure to contaminants (United Nations International Children’s Emergency Fund (UNICEF), 2025; WHO, 2017), and their developing physiological defense systems also make them more susceptible to irreversible damage from toxic exposures during this critical developmental window. Furthermore, young children’s behaviors, such as hand-to-mouth activity and proximity to ground-level pollutants, create unique exposure pathways (UNICEF, 2025; WHO, 2017). These early-life exposure can result in a range of adverse outcomes, such as impaired neurodevelopment, respiratory illness, and an increased risk of chronic diseases later in life (Landrigan et al., 2018; WHO, 2017).

In the U.S., health disparities are longstanding societal challenges, driven by a number of social determinants including race/ethnicity, income, education, house instability, access to healthcare, food insecurity etc. (Macias-Konstantopoulos et al., 2023). These inequities are also evident in children’s health, which is even more complex because children’s health outcomes emerge from interactions across multiple systems (Bronfenbrenner & Morris, 2007). At the microsystem level, children’s health is directly affected by immediate family and caregiving environment, such as the family’s ability to secure adequate shelter, nutrition, and healthcare. Exosystem factors such as community environment, extended families, and local industries, as well as broader societal forces at the macrosystem level, including economic policies, cultural norms, and systemic inequality, can also play a critical role in children’s health outcomes. When viewed through a bioecological lens (Bronfenbrenner & Morris, 2007), these different level influences converge most acutely in marginalized settings.

An important dimension of these disparities lies in children’s environmental health. Children in marginalized communities often experience disproportionate exposure to environmental hazards. They often face a “double burden” from both traditional hazards, like household air and water pollution, and modern hazards from industrialization and chemical contamination (Zajac et al., 2025). These cumulative exposures often result in high levels of outdoor air pollutants like PM_2.5_, NO_2_, and ozone, which significantly increase the risk of childhood asthma, wheezing, and other respiratory conditions (Achakulwisut et al., 2019; Great Britain. Committee on the Medical Effects of Air Pollutants., 1995). Thus, there is an urgent need to address such disparity in children’s environmental health, which requires the development of robust tools capable of systematically assessing and quantifying community vulnerability.

Although numerous metrics have been developed to quantify community vulnerability in public health, none of them explicitly address the unique risks of young children’s environmental health. The SVI, for example has been extensively used in disaster management and public health research, however, it mainly captures sociodemographic themes and provides limited quantification of environmental exposures. Several exposure-specific indices, such as National Risk Index (NRI) (Federal Emergency Management Agency (FEMA), 2021) which integrates social vulnerability with exposure to 18 natural hazards, Community Health Vulnerability Index (CHVI) (Rappold et al., 2017) which evaluates health risks from wildfire smoke and air pollution, and CVI (Tee Lewis et al., 2023) which integrate multiple exposure domains, were designed for the general population, with children remaining significantly underrepresented (Iliscupidez et al., 2025). Additionally, some of those metrics are limited to single exposures and lack a comprehensive view of cumulative environmental impacts.

Recently, a child-centered measurement, Child Opportunity Index (COI), was developed with health and environment considered as one of its three domain to capture overall developmental opportunities for children aged under 18 years (Noelke et al., 2025a). The health and environment domain includes four subdomains: pollution, healthy environments, safety-related resources, and health resources, and its validity has been demonstrated using 29 health and 7 socioeconomic outcome variables (Noelke et al., 2025a). While the COI highlighted the importance of recognizing children’s specific needs, it was designed for children in all age rather than young children under 5, the age stage of greatest physiological vulnerability. Moreover, it aims to provide a broad measure of opportunity rather than a targeted assessment of environmental health risks. Therefore, the environmental health disparity of young children, the group most physiologically and behaviorally susceptible, remain understudied.

To fill this gap, we developed of a county-level EC-EHVI for the contiguous U.S., with 2023 as the target year. Specifically, we adopted an Environmental Exposure, Sensitivity, and Adaptive Capacity framework and incorporated concepts from Bronfenbrenner’s bioecological model to capture multidimensional factors that might affect young children’s environmental health. We also examined the drivers (i.e., exposure-driven, sensitivity and adaptive capacity-driven, and co-driven) and spatial distributions of the EC-EHVI. This study can offer a comprehensive framework for assessing early childhood environmental health conditions and provides insights to help mitigate disparities across the U.S.

## 2 Methods

### 2.1 Study Area

This study includes all 3,109 counties in the 48 contiguous U.S. and the District of Columbia (CONUS). Alaska, Hawaii, and Puerto Rico were excluded to ensure data completeness and methodological consistency, as many national-level environmental and socio-economic indicators are either unavailable or reported using different methods for these regions (Centers for Disease Control and Prevention (CDC), 2024; U.S. Census Bureau, 2024; U.S. Environmental Protection Agency (EPA), 2016). We conducted the analyses at the county level, the scale at which reliable, multi-domain data are consistently available.

### 2.2 EC-EHVI Development and Validation

#### 2.2.1 Variable Selection

We selected indicators for the development of the EC-EHVI based on the Exposure-Sensitivity-Adaptive Capacity (ESA) framework and Bronfenbrenner’s bioecological model of human development (Bronfenbrenner, 2007; Intergovernmental Panel on Climate Change (IPCC), 2001; Smit et al., 2001). This dual conceptual foundation ensured that the variables capture not only the key functional dimensions of environmental health stressors but also bioecological factors that shape child development ranging from individual and family characteristics to broader community and societal contexts.

Based on this dual conceptual framework, each indicator was required to meet two criteria: (1) relevance, demonstrated through established importance in environmental health or community-level health research; and (2) availability, with standardized and comparable data accessible at the county level across the contiguous U.S. In total, 43 indicators were retained. These indicators are organized hierarchically into 16 subdomains, which are then grouped into three primary domains: Exposure, Sensitivity, and Adaptive Capacity. For each indicator, we primarily used the county-level estimate from 2023. When 2023 data were unavailable, we used the most temporally proximate county-level estimates to 2023. Three birth-related indicators (low birth weight, maternal age risk, and unmarried birth rate) were aggregated across 2019–2023 to reflect cumulative birth conditions for children aged 0 – 5 in 2023.

In the Exposure domain, we considered 14 indicators across four subdomains: air pollution, food and water, built and natural environment, and disaster and natural hazard risk (Table 1). On a per pound of body weight basis, children breathe far more air than adults, which results in them inhaling relatively higher doses of air pollutants (Jacobson & Wagner, 2024). Particulate matter, O3, NO2, SO2 and CO are recognized as the major air pollutants with the most significant adverse impacts on public health (WHO, 2021). Therefore, this study focuses on these five pollutants. In addition to air pollution, we also consider contaminated water sources and food safety, as exposure to heavy metals, nitrates, and pesticide residues can impair children’s organ development, disrupt endocrine and immune functions, and increase the risk of chronic diseases and developmental delays (Coleman-Jensen et al., 2023; UNICEF, 2025; WHO, 2017). Exposures from the built and natural environment, including noise pollution, road traffic density, impervious surface coverage, and tree canopy, which directly or indirectly affect children’s health, were also considered (Jacobs et al., 2007; Markevych et al., 2017). Furthermore, we included the National Hazard Risk Score from FEMA’s National Risk Index, which quantifies exposure to 18 natural hazards, such as avalanches, coastal flooding, heat waves, droughts, earthquakes, hurricanes, wildfires, and winter storms, among others (FEMA, 2022).

**Table 1.**
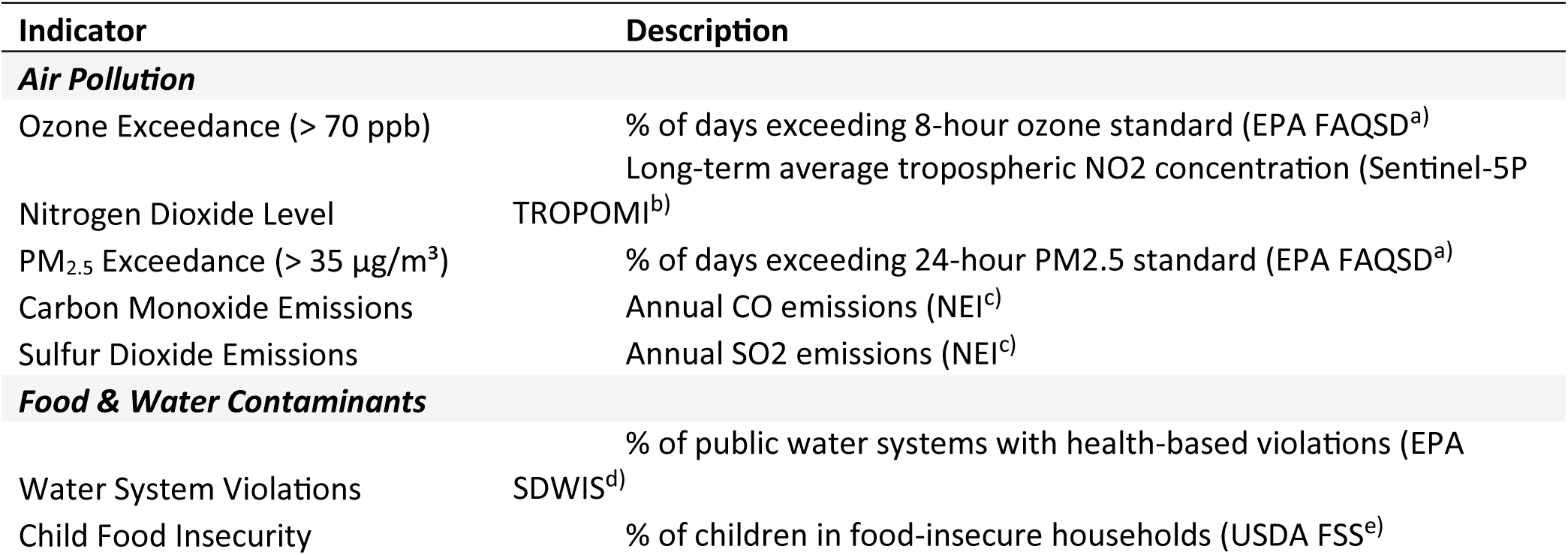
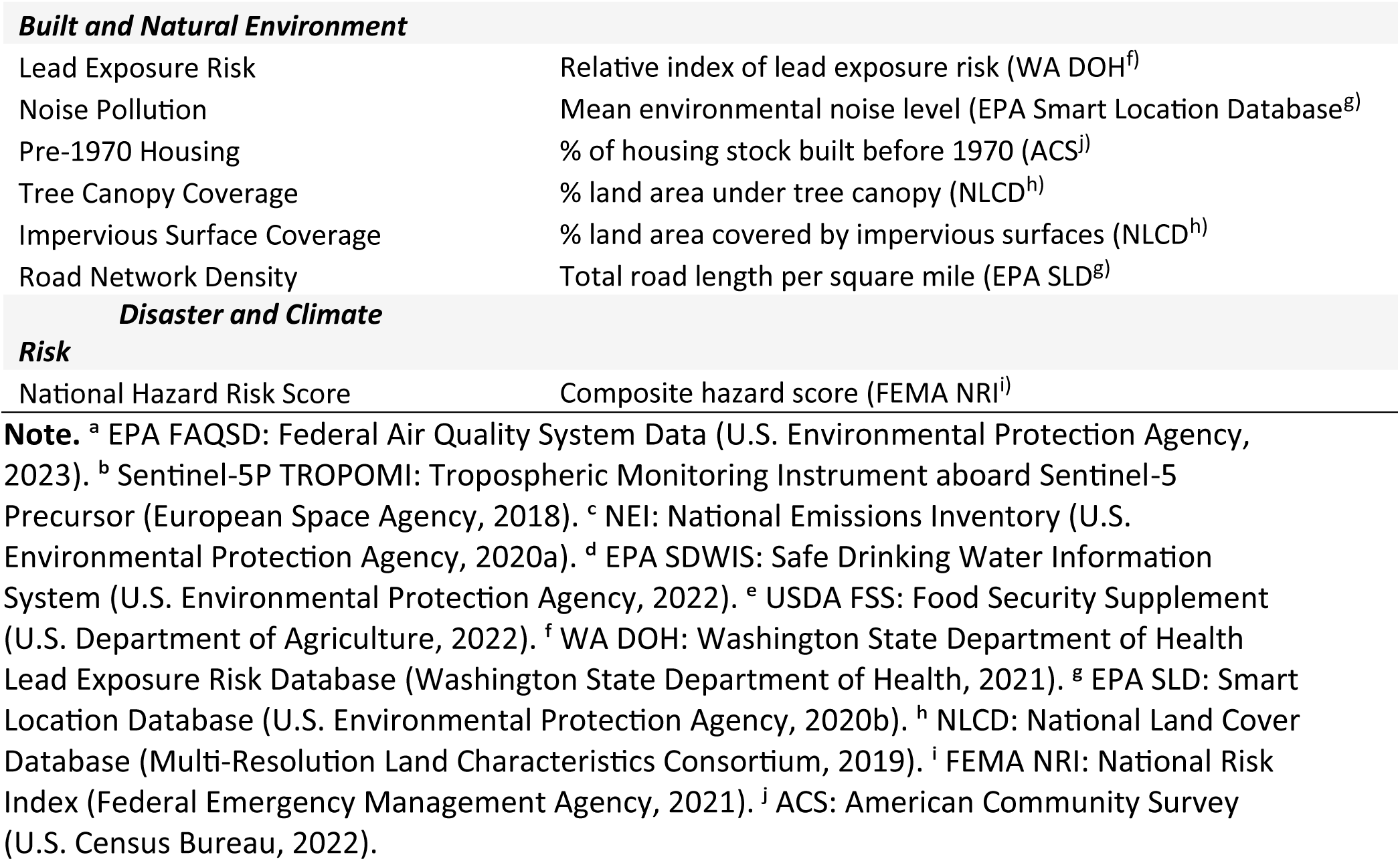
Environmental Exposure Domain Indicators.

**Table 2.** Health Sensitivity Domain Indicators.

| **Indicator** | **Description** |
| --- | --- |
| ***Individual Susceptibility*** |  |
| Low Birth Weight | % of live births under 2,500 grams (CHR<sup>a</sup>) |
| Maternal Age Risk | % births to mothers < 20 or > 35 (CHR<sup>a</sup>) |
| **Familial Health Susceptibility** |  |
| Obesity Prevalence | % adults (18+) with BMI ≥ 30 (CDC PLACES<sup>b</sup>) |
| Asthma Prevalence | % adults ever diagnosed with asthma (CDC PLACES<sup>b</sup>) |
| Cholesterol Screening Prevalence | % adults ever diagnosed with high cholesterol (CDC PLACES<sup>b</sup>) |
| Diabetes Prevalence | % adults diagnosed with diabetes (CDC PLACES<sup>b</sup>) |
| Hypertension Prevalence | % adults diagnosed with high blood pressure (CDC PLACES<sup>b</sup>) |
| **Family Structure** |  |
| Unmarried Birth Rate | % of live births to unmarried mothers (ACS<sup>c</sup>) |
| Single-Parent Households | % of children under 18 in single-parent households (ACS<sup>c</sup>) |
| **Community Health Susceptibility** |  |
| Physical Health (≥ 14 Days Poor) | % adults reporting ≥ 14 days poor physical health (CDC PLACES<sup>b</sup>) |
| Mental Health (≥ 14 Days Poor) | % adults reporting ≥ 14 days poor mental health (CDC PLACES<sup>b</sup>) |
| **Community Structure** |  |
| Labor Force Participation | % adults in labor force (ACS<sup>c</sup>) |
| Non-Hispanic White Population | % non-Hispanic White residents (ACS<sup>c</sup>) |
| Renter-Occupied Housing | % occupied housing rented (ACS<sup>c</sup>) |

**Note.** <sup>a</sup> CHR: County Health Rankings & Roadmaps (County Health Rankings, 2023). <sup>b</sup> CDC PLACES: Local Data for Better Health (U.S. Centers for Disease Control and Prevention, 2022). <sup>c</sup> ACS: American Community Survey (U.S. Census Bureau, 2022).

In the Sensitivity domain, we considered susceptibility at the individual, family, and community levels, using 14 indicators grouped into five subdomains (**Table2**). At the individual level, children’s health status was represented by indicators such as low birth weight and maternal age, both of which are well-documented predictors of early-life vulnerability (Aradhya et al., 2023). At the family level, we incorporated intergenerational health risks, including heritable or familial tendencies toward chronic conditions such as obesity, asthma, and cholesterol-related disorders (Khoury et al., 1993). Family structure is also closely linked to child health outcomes. For instance, children from single-parent or unmarried households often experience higher levels of economic insecurity, psychosocial stress, and reduced access to health-promoting resources, which may contribute to poorer physical and mental health outcomes (The Annie E. Casey Foundation, 2024). Accordingly, we included indicators of family structure such as the unmarried birth rate and the proportion of single-parent households. Finally, at the community level, we accounted for broader determinants of vulnerability, including overall physical and mental health prevalence, as well as structural characteristics of the community such as labor force participation, racial composition (non-Hispanic White population), and the proportion of renter-occupied housing, which together shape the social and economic environment influencing child health (Leventhal & Brooks-Gunn, 2000; Williams & Collins, 2001).

In the Adaptive Capacity domain, we included 15 indicators across seven subdomains, reflecting both resources relevant to environmental health adaptation and the accessibility of those resources (**Table 3**). Resources encompass economic conditions, healthcare, education, insurance, and policy supports, which are well-documented determinants of population health and resilience (Andersen, 1995; Cutler & Lleras-Muney, 2006; Kronick, 2013; Mande, 2023; Ross & Wu, 1995). Economic, educational, and insurance indicators were assessed at both child and adult levels to capture children’s conditions and their broader environments. Healthcare was assessed not only through hospital density, representing service availability, but also through rates of routine check-ups, which indicate preventive health awareness and utilization. Policy supports were represented by measures such as child tax credit coverage, public assistance, and food assistance programs. Accessibility of resources was represented by the Technology & Communication and Mobility subdomains, including household access to computers and telephones and the Neighborhood Walkability Index (Agency, 2014).

**Table 3.**
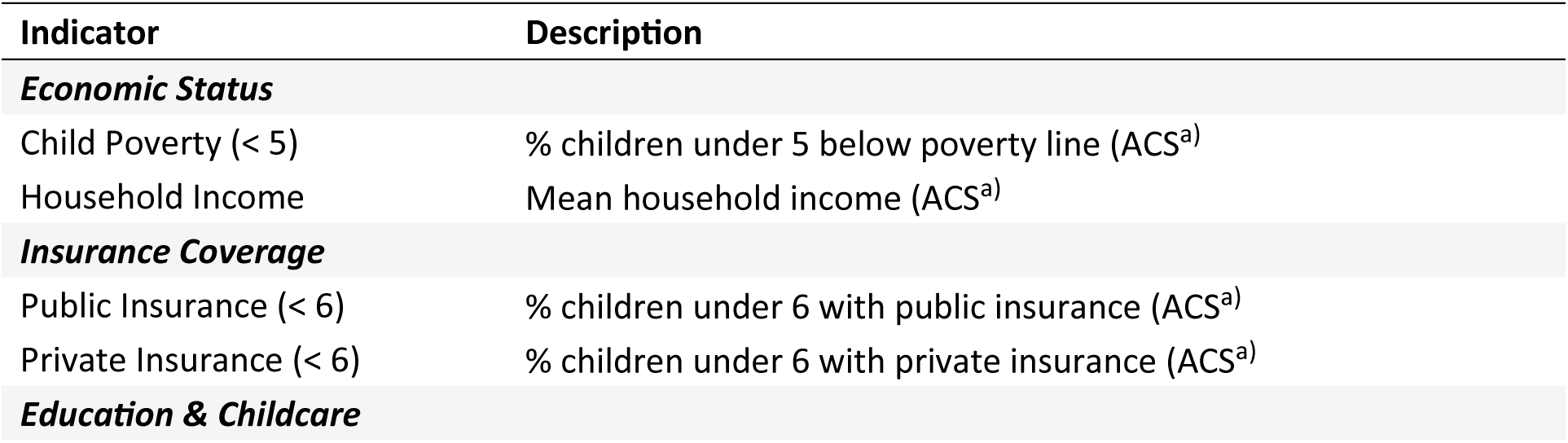
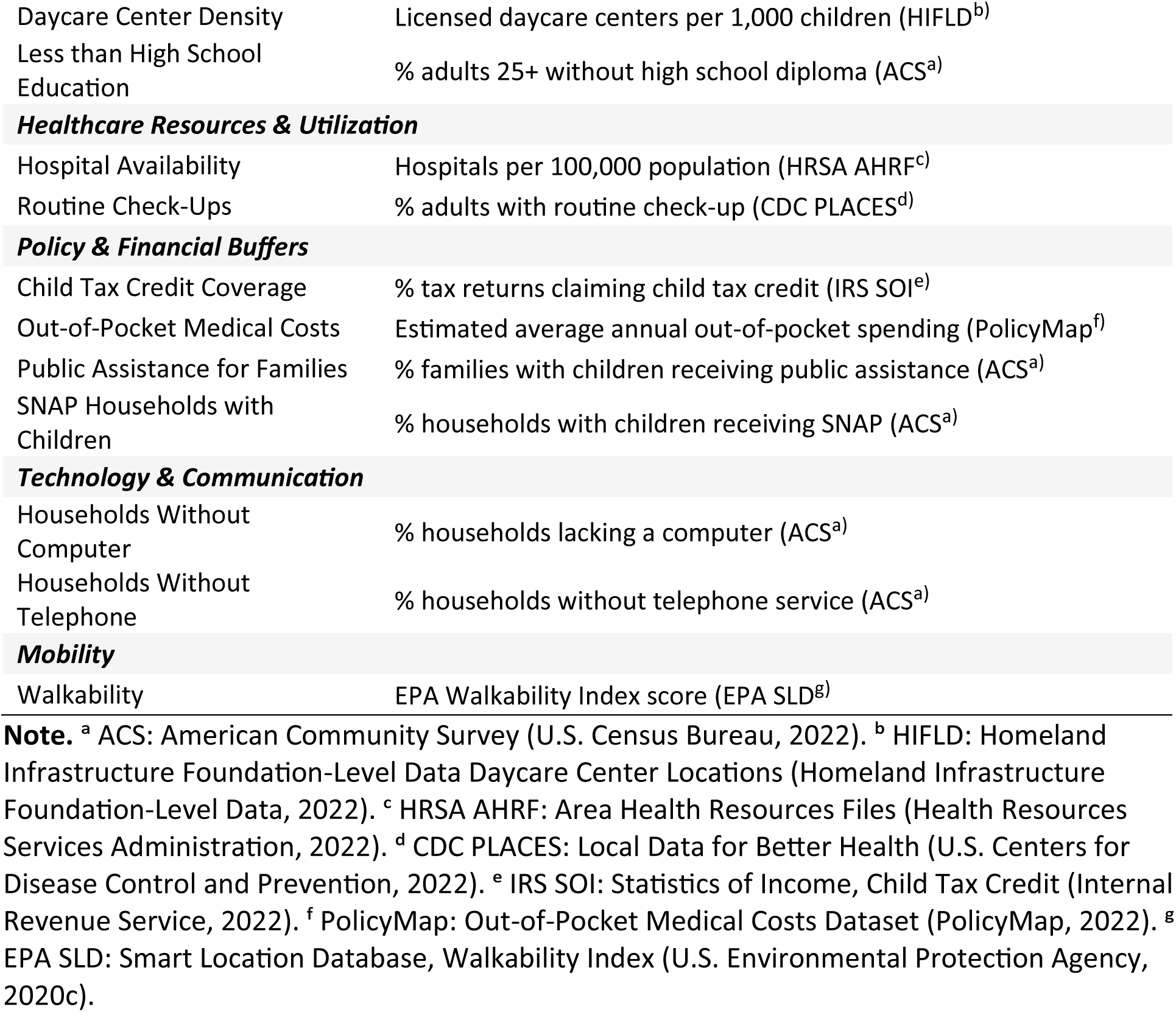
Adaptive Capacity Domain Indicators.

| Indicator | Description |
| --- | --- |
| **Economic Status** |  |
| Child Poverty (< 5) | % children under 5 below poverty line (ACS<sup>a</sup>) |
| Household Income | Mean household income (ACS<sup>a</sup>) |
| **Insurance Coverage** |  |
| Public Insurance (< 6) | % children under 6 with public insurance (ACS<sup>a</sup>) |
| Private Insurance (< 6) | % children under 6 with private insurance (ACS<sup>a</sup>) |
| **Education & Childcare** |  |
| Daycare Center Density | Licensed daycare centers per 1,000 children (HIFLD<sup>b</sup>) |
| Less than High School Education | % adults 25+ without high school diploma (ACS<sup>a</sup>) |
| **Healthcare Resources & Utilization** |  |
| Hospital Availability | Hospitals per 100,000 population (HRSA AHRF<sup>c</sup>) |
| Routine Check-Ups | % adults with routine check-up (CDC PLACES<sup>d</sup>) |
| **Policy & Financial Buffers** |  |
| Child Tax Credit Coverage | % tax returns claiming child tax credit (IRS SOI<sup>e</sup>) |
| Out-of-Pocket Medical Costs | Estimated average annual out-of-pocket spending (PolicyMap<sup>f</sup>) |
| Public Assistance for Families | % families with children receiving public assistance (ACS<sup>a</sup>) |
| SNAP Households with Children | % households with children receiving SNAP (ACS<sup>a</sup>) |
| **Technology & Communication** |  |
| Households Without Computer | % households lacking a computer (ACS<sup>a</sup>) |
| Households Without Telephone | % households without telephone service (ACS<sup>a</sup>) |
| **Mobility** |  |
| Walkability | EPA Walkability Index score (EPA SLD<sup>g</sup>) |

**Note.** <sup>a</sup> ACS: American Community Survey (U.S. Census Bureau, 2022). <sup>b</sup> HIFLD: Homeland Infrastructure Foundation-Level Data Daycare Center Locations (Homeland Infrastructure Foundation-Level Data, 2022). <sup>c</sup> HRSA AHRF: Area Health Resources Files (Health Resources Services Administration, 2022). <sup>d</sup> CDC PLACES: Local Data for Better Health (U.S. Centers for Disease Control and Prevention, 2022). <sup>e</sup> IRS SOI: Statistics of Income, Child Tax Credit (Internal Revenue Service, 2022). <sup>f</sup> PolicyMap: Out-of-Pocket Medical Costs Dataset (PolicyMap, 2022). <sup>g</sup> EPA SLD: Smart Location Database, Walkability Index (U.S. Environmental Protection Agency, 2020c).

All variables considered in those three domains were mapped out onto the hierarchical layers of Bronfenbrenner’s bioecological model to show how they collectively capture multi-level influences on early childhood environmental health and well-being. Specifically, based on this theoretical framework, we categorized the indicators into five distinct levels: Child; Parent & Family; Supportive School, Work, and Religious Settings; Neighborhoods and Communities; and Social, Economic, and Cultural Contexts and Policies (**Table 4**).

**Table 4.** Variables Mapped to the Bioecological Model Layers.

| Bioecological Layer | Variables |
| --- | --- |
| Child | Low Birthweight, Maternal Age |
| Parent & family | Unmarried Birth Rate, Single-Parent Households, Obesity Prevalence, Asthma Prevalence, Cholesterol Screening Prevalence, Diabetes Prevalence, Hypertension Prevalence, Public Insurance (< 6), Private Insurance (< 6) |
| Supportive school, work, and religious settings | Daycare Center Density, Routine Check-Ups, Hospital Availability, Less than High School Education |
| Neighborhoods and communities | Ozone Exceedance, Nitrogen Dioxide Level, PM<sub>2.5</sub> Exceedance, Carbon Monoxide Emissions, Sulfur Dioxide Emissions, Water System Violations, Child Food Insecurity, Lead Exposure Risk, Noise Pollution, Pre-1970 Housing, Tree Canopy Coverage, Impervious Surface Coverage, Road Network Density, National Hazard Risk Score, Labor Force Participation, Non-Hispanic White Population, Renter-Occupied Housing, Walkability |
| Social, economic and cultural contexts and policies | Child Poverty (< 5), Household Income, Households Without Computer, Households Without Telephone, Child Tax Credit Coverage, Out-of-Pocket Medical Costs, Public Assistance for Families, SNAP Households with Children |

#### 2.2.2 Index Construction

Before the construction of the EC-EHVI, we winsorized each indicator at the 1^st^ and 99^th^ percentiles to mitigate the influence of extreme outliers while retaining all counties. We then harmonized the indicator directionality by inverting variables where lower scores denoted higher concern, ensuring that a higher score consistently represented higher environmental health vulnerability (**Table S1**). Missing values were imputed using the K-nearest neighbor (KNN) algorithm. The optimal neighborhood size (K) in the KNN algorithm was determined by systematically testing integer values from 1 to 20, a commonly recommended range for balancing bias and variance (James et al., 2021), and selecting the value that minimized the Root Mean Square Error (RMSE) on a stratified random subset of the data (n = 1,000), which resulted in K = 5. Finally, all variables were standardized using z-scores to ensure a uniform scale prior to index construction.

The EC-EHVI was constructed using a hierarchical Multi-Criteria Evaluation (MCE) approach with equal weighting. This equal-weighting approach was intentionally chosen to preserve the conceptual balance of the theoretical framework. It also avoids the introduction of subjective or complex data-driven weights that could obscure the distinct contributions of each component to overall vulnerability. Equal weighting was applied at each aggregation level, from indicator to the subdomain and then to the primary domain. This process began with the 43 standardized indicators, which were averaged to generate scores for 16 subdomains. These subdomain scores were then aggregated to produce scores for the three primary domains of Exposure, Sensitivity, and Adaptive Capacity. After the establishment of the index structure, we examined the spatial distribution of vulnerability at county level using both continuous scores and quintile classification. To identify broader regional trends, county-level scores were then averaged to the state level. Finally, we summarized the statistical distributions of the EC-EHVI and its domains by computing the mean, standard deviation, and interquartile range.

#### 2.2.3 Validation and Sensitivity Analysis

We employed both predictive and convergent approaches to validate the EC-EHVI. For predictive validation, we assessed the performance of the EC-EHVI in explaining county-level all-cause early childhood mortality counts in 2023, with population size accounted for through an offset term, and compared its performance with that of existing indices (Centers for Disease, Prevention, & National Center for Health, 2024). Given the significant spatial autocorrelation within the mortality rates (Global Moran’s I = 0.42, p < 0.001), we employed a Bayesian Spatial Model using the Besag-York-Mollié (BYM2) framework (Banerjee et al., 2004; Besag et al., 1991; Lindgren et al., 2011; R Core Team, 2025; Rue et al., 2022; Rue et al., 2009), which can capture both structured spatial dependencies and unstructured random noise. We specified a Negative Binomial likelihood to accommodate overdispersion in mortality counts and included the log of the county-level early childhood population as an offset, thereby modeling mortality risk while accounting for population size. The Bayesian parameters were estimated with the Integrated Nested Laplace Approximation (INLA).

We then compared EC-EHVI performance against other four existing indices: SVI, COI, the COI’s health and environment domain (COI-HE), and CVI (Centers for Disease, Prevention, Agency for Toxic, et al., 2024; Lewis et al., 2023; Noelke et al., 2025b). Associations between each vulnerability index and the outcome were estimated using relative risk (RR) estimates with 95% credible intervals (CrI), scaled per one-standard deviation increase in each index. Model performance was evaluated using the Watanabe-Akaike Information Criterion (WAIC), with lower values indicating better overall model fit. Finally, the ability of each index to account for spatial heterogeneity was assessed by examining the total random effect variance from the spatial model.

For convergent validation, we examined the consistency and robustness of EC-EHVI with existing indices through Spearman’s rank correlations at the county level. We interpreted the correlation strength based on established guidelines: weak (|r_s_|: 0.20 – 0.39), moderate (0.40 – 0.59), and strong (≥ 0.60) (Evans, 1996).

We also conducted the sensitivity analyses to evaluate the stability of EC-EHVI by reconstructing the index under alternative preprocessing choices, including no outlier treatment and ± 3 SD removal, as well as Min–Max normalization, and comparing the results with the preprocessing approach used in this study (1% winsorization and Z-score standardization). Additionally, we also compared two alternative index construction frameworks: a direct Equal-Weighting of all 43 indicators and a Principal Component Analysis (PCA) derived score. The robustness of our final model was evaluated by assessing the consistency of county-level rankings and statistical associations across these different methodological specifications.

### 2.3 Drivers of Vulnerability

To identify the dominant drivers of vulnerability, each county was categorized into one of three groups: exposure-driven, sensitivity and adaptive capacity-driven, or co-driven. A county was classified as exposure-driven when its standardized exposure score exceeded the mean of its sensitivity and adaptive capacity scores by at least 0.5 standard deviations (SD). Conversely, a county was classified as sensitivity/adaptive capacity-driven when the mean of its sensitivity and adaptive capacity scores was ≥ 0.5 SD higher than the exposure score. Counties with absolute differences < 0.5 SD were considered co-driven, indicating a relatively balanced influence from both domains. The 0.5 SD threshold was selected following common practices for identifying moderate effect sizes and meaningful differentiation in standardized indices (Cohen, 2013).

This classification simplifies the ESA framework by combining Sensitivity and Adaptive capacity into a composite dimension. The decision was supported by both empirical and conceptual considerations. Empirically, correlation analysis indicated strong overlap between sensitivity and adaptive capacity (Spearman’s r > 0.8; **Table S2**), suggesting redundancy if treated separately. Conceptually, interventions aimed at reducing social sensitivity, such as increasing income levels, improving education, and enhancing housing stability, often simultaneously strengthen adaptive capacity by promoting community resources and institutional resilience. In contrast, interventions targeting exposure, such as air-quality control, urban greening, and climate mitigation, primarily operate within the environmental domain and have limited direct influence on socioeconomic resilience. Distinguishing between environmental (E) and socioeconomic ((S + A) / 2) drivers thus yields a more interpretable and application-relevant framework.

To ensure robustness, we tested an alternative threshold of 1 SD. This stricter rule classified more counties as co-driven but did not alter the overall spatial pattern of results (**Table S3** and **Figure S1**). The chosen 0.5 SD threshold, therefore, provides a balance between sensitivity and parsimony.

### 2.4 Spatial Analysis of EC-EHVI

We conducted a two-stage analysis to formally assess the geographic distribution of EC-EHVI. First, we calculated the Global Moran’s I statistic to determine if the EC-EHVI and its components exhibited significant spatial clustering across the entire study area. We then employed two complementary local methods to identify the location of these clusters if the significant global clustering was identified. Specifically, we employed Local Indicators of Spatial Association (LISA) to detect significant spatial patterns, including high–high clusters (hot spots), low–low clusters (cold spots), and spatial outliers. In parallel, the Getis-Ord Gi* statistic was used to assess the intensity and statistical confidence of clustering, with significance evaluated at the 90%, 95%, and 99% levels. All spatial autocorrelation analyses were based on a first-order queen contiguity spatial weights matrix, and statistical significance was determined using 9,999 Monte Carlo permutations.

To increase robustness, we focused on double-identified clusters, defined as counties simultaneously classified as high-score clusters by both LISA (p < 0.05) and Gi* (Z-score > 1.96). Within these clusters, we further examined the distribution of driver types. This step links spatial clustering patterns with their underlying drivers, enabling interpretation of whether geographically entrenched risks are primarily environmental, socioeconomic, or compounded.

Spatial analyses were conducted using the spdep package (for LISA and Getis-Ord Gi* analyses), sf package (for spatial data handling), and ggplot2 with ggspatial (for map visualization and annotation) (Bivand et al., 2013; Bivand & Wong, 2018; Dunnington, 2021; Pebesma, 2018; R Core Team, 2025; Wickham, 2016).

## 3 Results

### 3.1 EC-EHVI Development and Validation

#### 3.1.1 Descriptive Patterns of EC-EHVI

We visualized the spatial distribution of EC-EHVI in both the continuous map to show gradient variation and the classified map to highlight distinct spatial patterns (**Figure 1**). Both representations demonstrate pronounced spatial disparities in environmental health vulnerability for early childhood.

**Figure 1.**
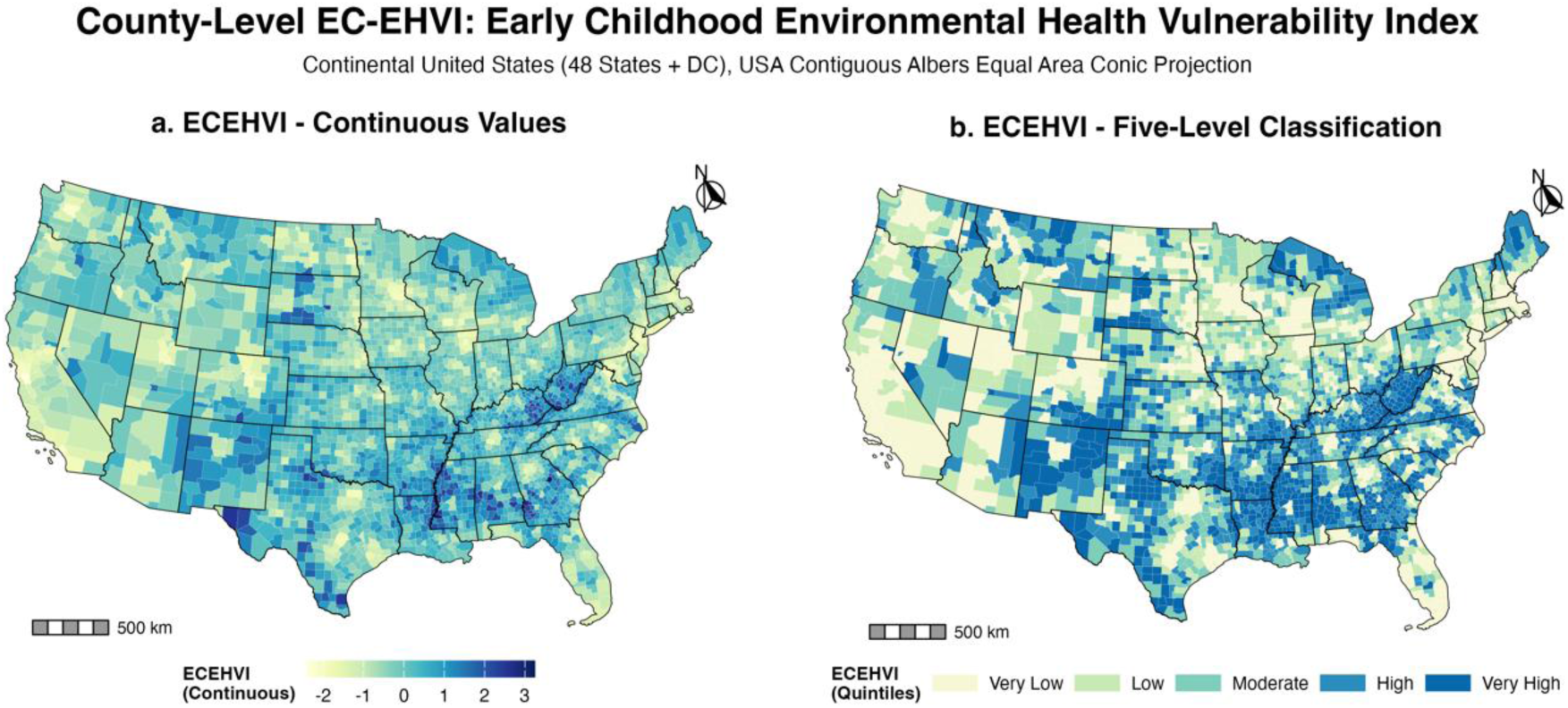
Spatial Distribution of the Early Childhood Environmental Health Vulnerability Index (EC-EHVI). The figure displays the EC-EHVI for 3,109 counties in the contiguous U.S. (a) Continuous EC-EHVI scores, where higher values (progressing from yellow to dark purple) indicate greater environmental health vulnerability. (b) EC-EHVI scores classified into quintiles, highlighting counties in the highest (Very High) and lowest (Very Low) 20% of environmental health vulnerability.

Overall, the map reveals a broad south–north gradient. Counties with lower vulnerability index scores are predominantly concentrated in the Northern and Western U.S. In contrast, counties with higher vulnerability index scores exhibit two prominent spatial patterns: (1) a contiguous north-south band extending from the Southern Great Plains and New Mexico northward through Nebraska and South Dakota to Montana, and (2) a large, densely clustered region encompassing the Southeastern U.S. and Appalachia. Additional clusters of high vulnerability areas are evident in Michigan and Maine.

The disaggregated component maps (**Figure 2**) showed how the spatial distribution of each component of Sensitivity, Exposure, and Adaptive Capacity shapes the composite index. The Exposure domain shows high scores in the Great Plains and western U.S., reflecting localized source of pollution and environmental stressors. The Sensitivity domain largely mirrors the distribution of the composite EC-EHVI, with high scores in the south, which is a known landscape of adverse health and social challenges. The Adaptive Capacity domain exhibits lower scores in major metropolitan areas and economically robust regions, indicating greater capacity to respond to environmental stress.

**Figure 2.**
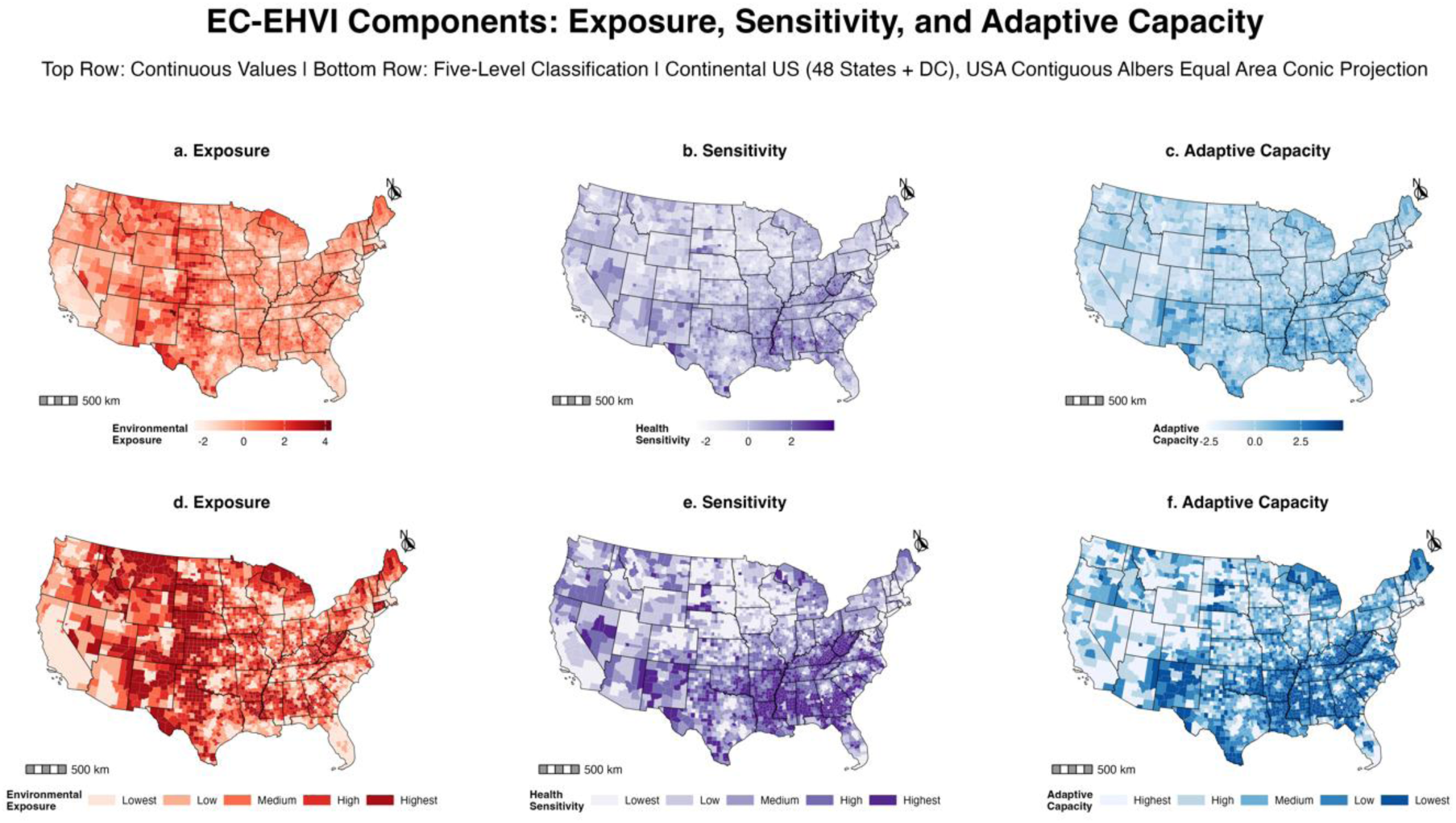
Spatial Distribution of the Three Core Components of the EC-EHVI. The figure displays the three component indices for 3,109 counties in the contiguous U.S., with continuous values in the top row (a-c) and quintile classifications in the bottom row (d-f). (a, d) Exposure: higher values (darker red) indicate greater environmental stressors. (b, e) Sensitivity: higher values (darker purple) indicate greater susceptibility to health impacts. (c, f) Adaptive Capacity: lower values (lighter blue) indicate greater adaptive capacity.

**Figure 3** showed distribution of the state level EC-EHVI scores averaged from county-level scores. A prominent concentration of states with high vulnerability emerges in the southeast, including ten states, i.e., West Virginia, Mississippi, Arkansas, Kentucky, Alabama, Louisiana, Georgia, New Mexico, Tennessee, and Oklahoma. In contrast, states with more favorable environmental health conditions for young children are predominantly clustered in the west (e.g., California and Utah) and the upper center (e.g., Minnesota and Wisconsin), with several states in the northeast (e.g., Massachusetts, Rhode Island, and New Jersey) also falling into this category.

**Figure 3.**
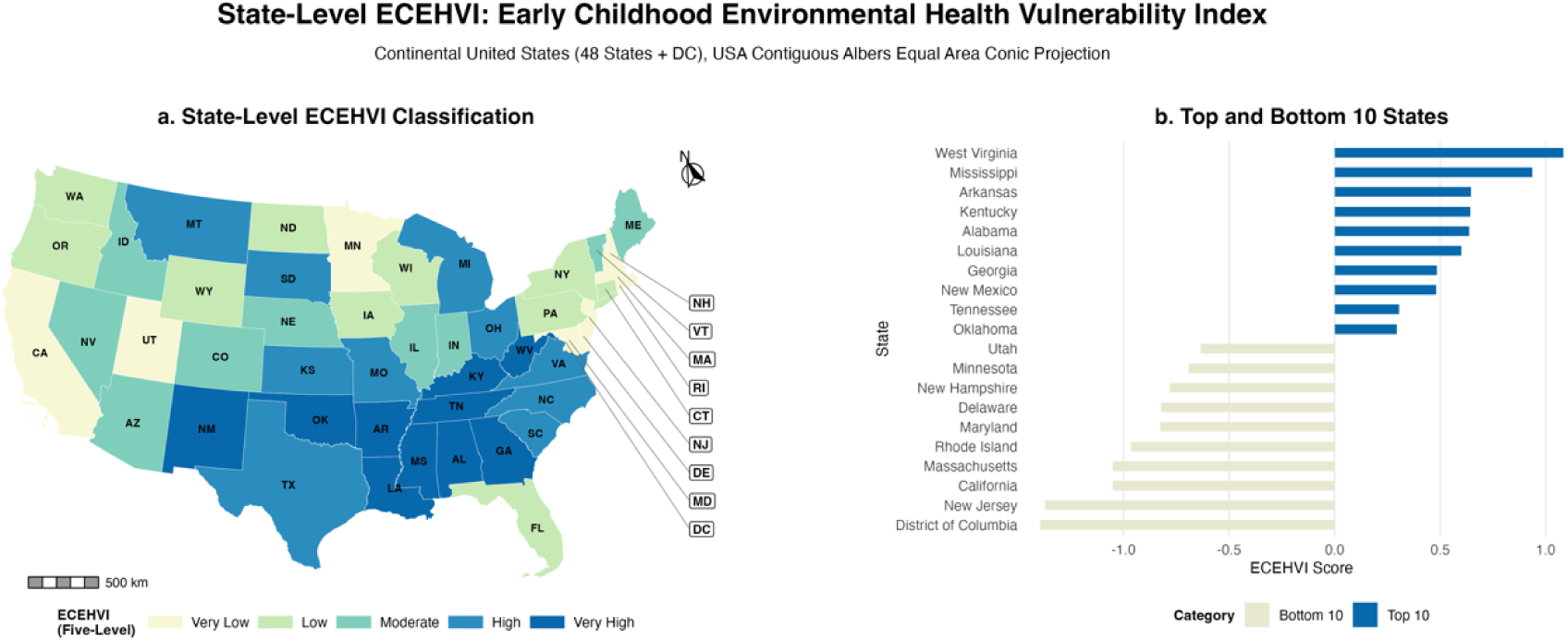
State-Level Patterns of the Early Childhood Environmental Health Vulnerability Index (EC-EHVI). (a) Choropleth map of state-level EC-EHVI scores, classified into five vulnerability levels. (b) Bar chart ranking the ten states with the highest vulnerability (purple) and the ten states with the lowest vulnerability (yellow).

#### 3.1.2 Validation

**Table 5** summarizes model performance and effect estimates for early childhood mortality across different vulnerability indices. Overall, the EC-EHVI demonstrated a strong and statistically robust association with early childhood mortality, with a 22% increase in risk per standard deviation increase (RR = 1.22, 95% CrI: 1.21 – 1.23). This effect size was notably larger than that observed for the SVI (RR = 1.03) and CVI (RR = 1.13), and opposite in direction to the COI-based indices. In terms of model fit, the EC-EHVI achieved a WAIC (33,020.00) that was lower than most alternative indices, although the CVI yielded the lowest WAIC overall, indicating good overall explanatory performance. Moreover, the EC-EHVI model exhibited the lowest total random effect variance (0.032), suggesting that it captured a larger proportion of spatially structured and unstructured heterogeneity in early childhood mortality relative to the other indices.

**Table 5.** Model performance and effect estimates in relation to early childhood mortality.

| Index | RR per SD (95% CrI) | Total RE Variance | WAIC |
| --- | --- | --- | --- |
| EC-EHVI | 1.22 (1.21 – 1.23) | 0.032 | 33020.00 |
| SVI | 1.03 (1.02 – 1.04) | 0.082 | 33295.10 |
| COI | 0.86 (0.85 – 0.86) | 0.050 | 33267.80 |
| COI-HE | 0.87 (0.86 – 0.88) | 0.060 | 33437.20 |
| CVI | 1.13 (1.12 – 1.15) | 0.078 | 32572.10 |

#### 3.1.3 Sensitivity Analysis

The EC-EHVI showed high robustness to data processing strategies, including alternative treatments of outliers and standardization methods. Across all specifications, relative risks (RRs) per standard deviation increase remained highly consistent (1.173 – 1.234), and Spearman’s rank correlations with the baseline hierarchical model consistently exceeded 0.99, indicating that these technical choices did not materially affect county-level rankings (**Table S4** and **Table S5**).

In terms of index construction approaches, the Equal-Weight and Hierarchical models performed comparably, and both significantly outperformed the PCA-based specification (**Table 6**). Although the Equal-Weight model achieved a marginally lower WAIC and higher RR per SD, the Hierarchical framework was retained as the primary model. This decision was based on its theoretical alignment with the ESA framework, its balanced domain weighting, and enhanced interpretability. Furthermore, the high correlation (Spearman’s ρ = 0.994) and highly similar RRs between the two models confirm that this choice does not compromise statistical robustness.

**Table 6.** Performance of Alternative Aggregation Methodologies.

| Methods | RR per SD (95% CrI) | Total RE Variance | WAIC |
| --- | --- | --- | --- |
| Final Model (MCE) | 1.22 (1.21 – 1.23) | 0.032 | 33020.00 |
| Equal Weighting | 1.23 (1.22 – 1.24) | 0.032 | 32930.00 |
| PCA Derived | 1.17 (1.16 – 1.19) | 0.051 | 33413.60 |

### 3.2 Primary Drivers of Vulnerability

We found that the primary driver of EC-EHVI scores was exposure in 30.5% of U.S. counties and sensitivity and adaptive capacity factors in 37%. For the remaining 32.6% of counties, both factors jointly determined the EC-EHVI (Figure 4). Spatially, exposure-driven counties largely overlapped with the high-vulnerability regions identified in Section 3.1.1. Specifically, exposure was the dominant factor in the north-south band (from New Mexico to Montana), the Southeastern US, Appalachia, and parts of Michigan and Maine. A notable exception was Iowa, where 38.4% of counties were exposure-driven despite exhibiting generally low overall EC-EHVI scores (mean value: – 0.383, rank 33/49, low vulnerability level). In contrast, the Western U.S. was predominantly characterized by sensitivity and adaptive capacity-driven patterns, while the Eastern U.S. displayed a heterogeneous mixture of co-driven and sensitivity and adaptive capacity-driven counties.

**Figure 4.**
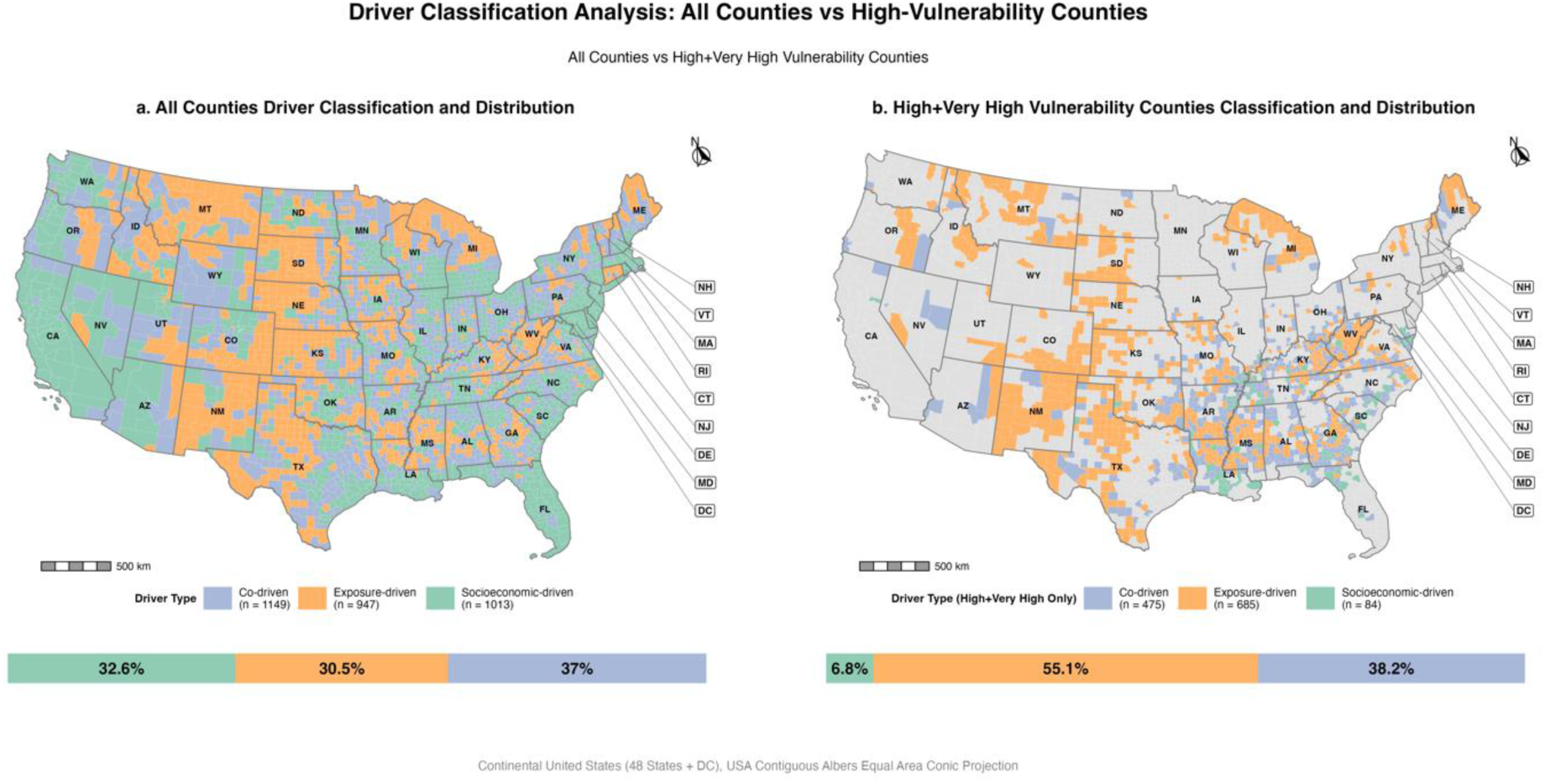
Driver Classification Analysis: All Counties vs High-Vulnerability Counties. (a) Choropleth map showing the dominant vulnerability driver types across all U.S. counties, classified into three categories: exposure-driven (orange), sensitivity and adaptive capacity-driven (green), and co-driven (blue), based on the relative difference in standardized component scores. (b) Choropleth map displaying driver type classification limited to counties identified as high and very high vulnerability (60^th^ – 80^th^ and > 80^th^ percentiles of EC-EHVI scores). The bar charts below each map summarize the proportion of counties by driver type in each group.

Among counties with High (60^th^ – 80^th^ percentiles) and Very High (> 80^th^ percentile) EC-EHVI scores, the majority (55.1%) were exposure-driven, followed by co-driven (38.2%), whereas sensitivity and adaptive capacity-driven counties comprised only 6.8%. This 55.1% share represents a substantial increase compared to the national baseline of 30.5%. It contrasts sharply with the overall pattern, where exposure is less dominant. Furthermore, a transitional spatial gradient was evident in **Figures 4a and 4b**. Exposure-driven counties were frequently bordered by co-driven areas, which subsequently transitioned into regions dominated by sensitivity and adaptive capacity.

### 3.3 Spatial Clustering and Shared Drivers

The Global Moran’s I analysis confirmed significant positive spatial autocorrelation for the EC-EHVI at the national scale (Global Moran’s I = 0.65, p < 0.001), indicating that high- and low-value counties tend to be geographically clustered, not randomly distributed. From **Figure 5**, a substantial portion of U.S. counties fall into statistically significant clusters (**Figure 5a**): Approximately 13.2% of counties formed statistically significant high-risk clusters and 13.2% formed low-risk clusters in the LISA analysis, while the Getis-Ord Gi* analysis similarly identified 18.0% of counties as high-score clusters and 18.5% as low-score clusters.

**Figure 5.**
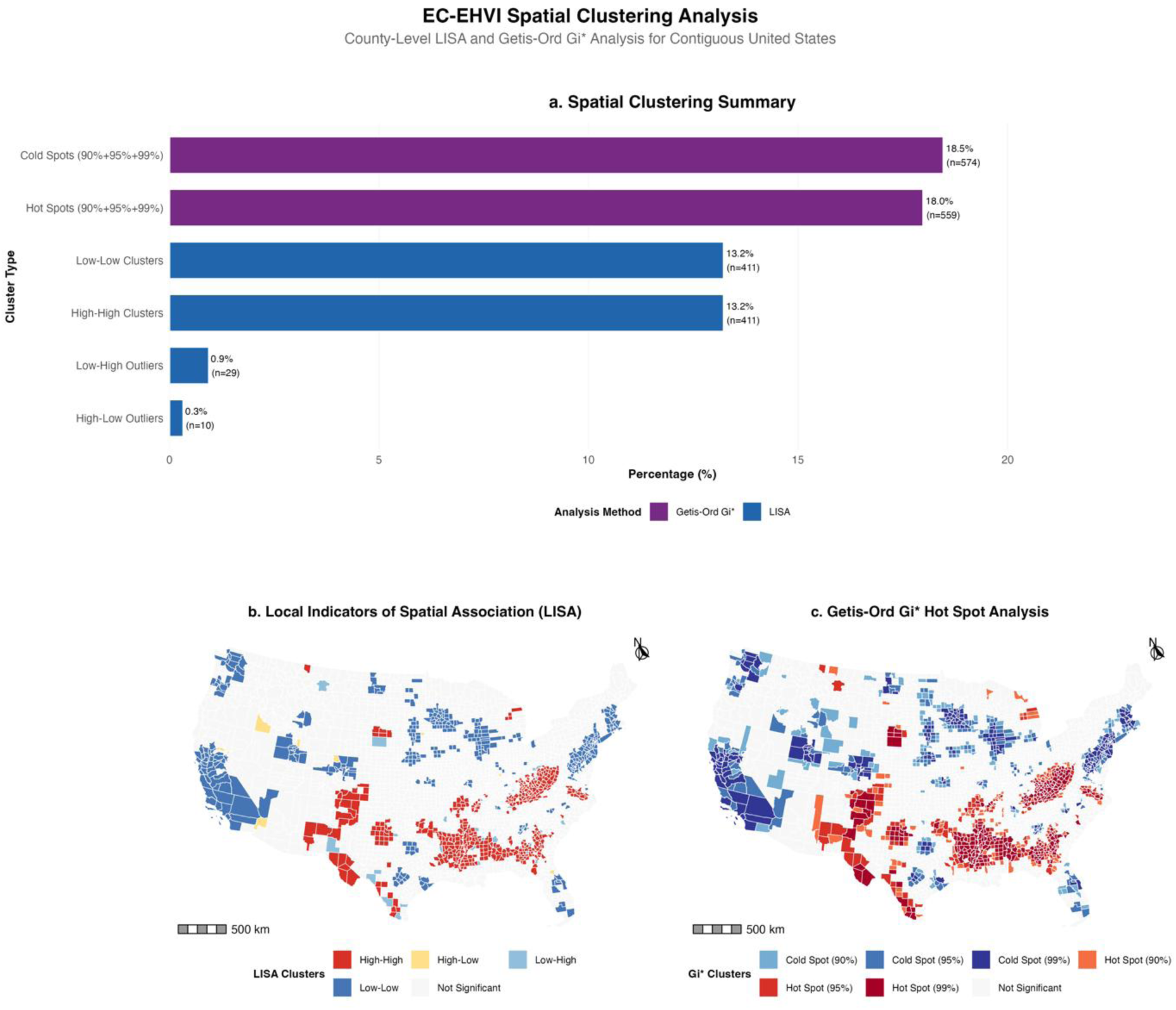
Spatial Clustering Analysis of the County-Level Early Childhood Environmental Health Vulnerability Index (EC-EHVI) using LISA and Getis-Ord Gi* statistics. (a) A summary of spatial clustering results, showing the percentage and count of counties within each cluster type identified by the two complementary analytical methods. (b) The geographic distribution of Local Indicators of Spatial Association (LISA) clusters, mapping significant High-High (vulnerability hot spots) and Low-Low (cold spots) clusters, as well as High-Low and Low-High spatial outliers. (c) The geographic distribution of Getis-Ord Gi* hot and cold spots, with results classified by their statistical significance (90%, 95%, or 99% confidence). Both analyses are based on a first-order queen contiguity spatial weights matrix.

The geographic distribution of these clusters is highly segregated (**Figure 5b – c**). A large, contiguous hot spot of high vulnerability dominates the Southern Great Plains, Southwest, and Appalachia. Conversely, three major Cold Spots, indicating more favorable environmental health conditions, are located in the northern U.S. The spatial distributions of the clusters were found to be overwhelmingly contiguous, with spatial outliers (High-Low and Low-High clusters) accounting for only approximately 1% of counties.

Using both the LISA and Gi* methods, we identified 411 counties across the continental U.S. that were consistently classified as high–high clusters (p < 0.05; Z-score > 1.96). Within these clusters, exposure-driven counties dominated (n = 245, 59.6%), followed by co-driven (n = 140, 34.1%) and sensitivity and adaptive capacity-driven counties (n = 26, 6.3%) (**Figure 6**).

**Figure 6.**
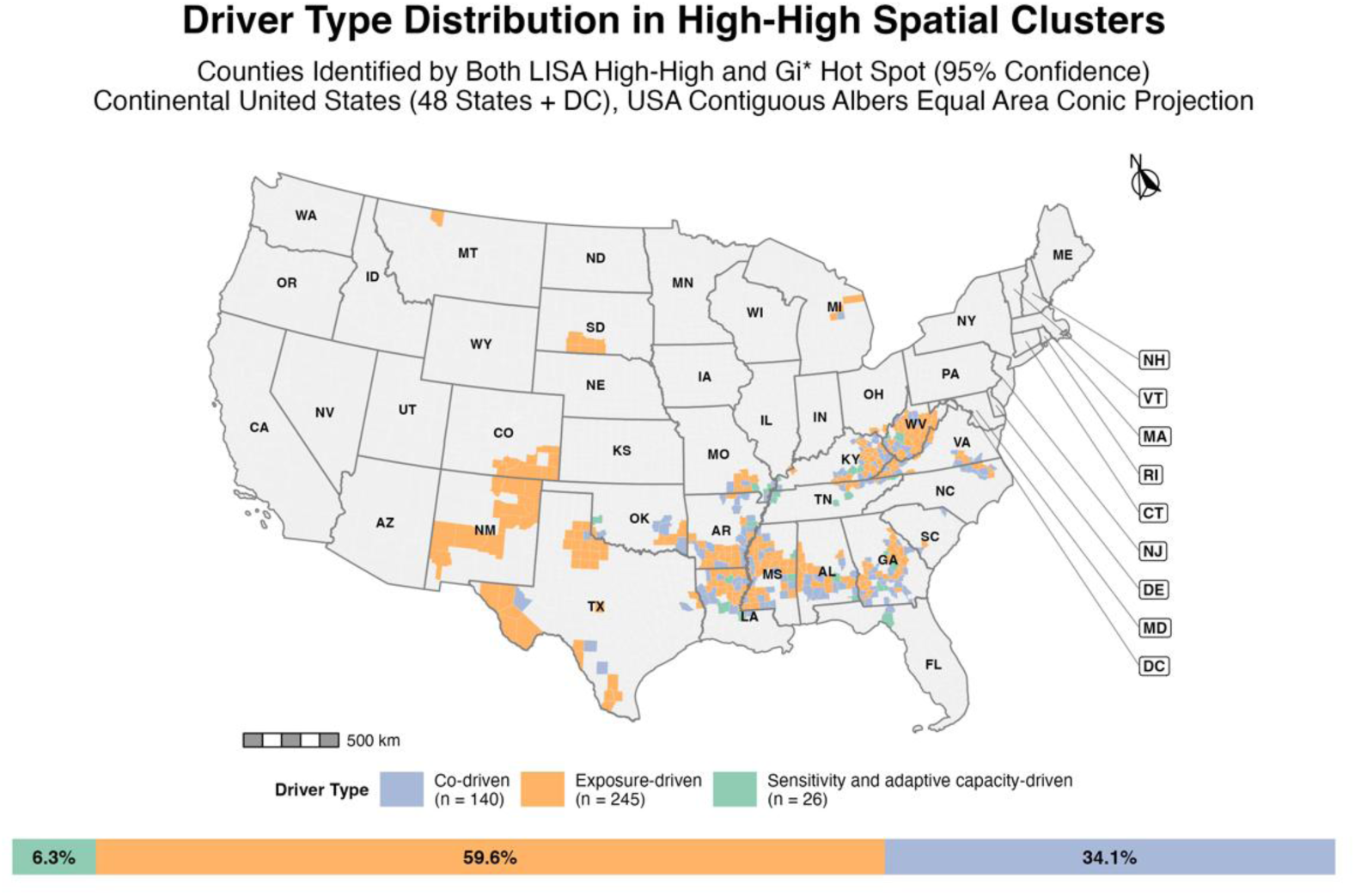
Distribution of dominant driver types of vulnerability across the 411 identified counties. The majority of clusters were exposure-driven (59.6%), followed by co-driven (34.1%), with Sensitivity and adaptive capacity-driven clusters (6.3%) being comparatively rare.

Spatially, exposure-driven clusters were primarily concentrated in the Southern Great Plains, Southwest, and Appalachia (e.g., Texas, New Mexico, Kentucky, and West Virginia). In contrast, co-driven and sensitivity and adaptive capacity-driven clusters were sparsely distributed, appearing mainly in the Southeast and often spatially adjacent to exposure-driven areas.

## 4. Discussion

Early childhood is a critical developmental period during which environmental exposures can create long-lasting health inequities, motivating this study to develop and validate the EC-EHVI to quantify these environmental health burdens across the contiguous U.S. The index revealed that counties with high and very high vulnerability were disproportionately concentrated in the north-south band (from New Mexico to Montana), the Southeastern U.S., and Appalachia, alongside distinct high-vulnerability clusters in Michigan and Maine. The EC-EHVI follows a spatial pattern similar to the COI Health and Environment domain, while also reflecting the north-south disparity also captured by other indices (Centers for Disease, Prevention, Agency for Toxic, et al., 2024; Diversitydatakids.org, 2025; FEMA, 2021).Importantly, in high vulnerability counties, more than half of these counties were exposure-driven, underscoring the dominant role of environmental hazards such as extreme heat and air pollution in shaping early childhood vulnerability. Furthermore, spatial clustering analysis confirmed that these risks are not randomly distributed but exhibit significant high–high clusters that remain predominantly exposure-driven, suggesting that environmental exposure burdens are spatially entrenched rather than isolated occurrences.

Although the CVI achieved the lowest WAIC among all models, indicating slightly better overall fit, several additional performance metrics highlight the added value of the EC-EHVI. Notably, the EC-EHVI exhibited the strongest positive association with early childhood mortality, with a substantially larger effect size per standard deviation compared to the CVI and SVI. Furthermore, the EC-EHVI model was associated with the smallest total random effect variance, suggesting that it explained a greater proportion of spatial heterogeneity in mortality risk rather than leaving it to unobserved area-level effects.

These performance advantages likely arose because the EC-EHVI was specifically designed to reflect young children’s environmental health vulnerability by integrating the ESA model and the bioecological model. This integration comprehensively captures the entire pathway from environmental exposure to child health outcomes. In contrast, widely used indices such as the SVI and CVI focus primarily on general population-level socioeconomic vulnerability or exclusively on environmental exposures, consequently overlooking the unique developmental and physiological vulnerabilities inherent to early childhood. Although newer indices like the COI and its Health & Environment domain incorporate child-related indicators, they are designed for the broader 0 – 18 population and still rely on a relatively limited set of environmental measures. Several important early-childhood exposures remain underrepresented.

Specifically, the EC-EHVI improves explanatory performance in three critical dimensions. First, the EC-EHVI integrates a more comprehensive set of environmental exposures than existing indices. For example, we captured five air pollutants while existing indices often restrict their focus to PM2.5 and ozone. We also incorporate noise, lead, and nature hazards that disproportionately affect young children but are frequently overlooked by existing indices. Second, we included as many as possible variables related to young children birth to age 5. Factors such as low birthweight and maternal age represent early-life biological sensitivity, while other child-specific variables such as daycare density, child private insurance, and child poverty capture social and economic conditions they face. Third, the index incorporated young child- and policy-relevant indicators including Child Tax Credit coverage, out-of-pocket medical costs, public assistance for families, and Supplemental Nutrition Assistance Program (SNAP) households with children. These reflect the institutional and structural supports but are typically overlooked in existing indices.

Overall, the spatial distribution of the EC-EHVI is most similar to the COI because both indices incorporate environmental and socioeconomic indicators relevant to children. We and the COI highlight overlapping areas of elevated vulnerability across the central and southwestern U.S. However, the EC-EHVI also diverges from the COI in several counties. These differences occur because the EC-EHVI focuses specifically on early-childhood environmental health risks, whereas the COI provides a broader assessment of developmental opportunity for the full 0 – 18 age range. Consequently, hazards that disproportionately affect young children receive greater weight in the EC-EHVI. For example, Wallace County in Kansas and Lexington City in Virginia are classified as High Opportunity areas by the COI (Diversitydatakids.org, 2025), yet our index identifies them as highly vulnerable due to severe drinking-water violations in Wallace County and elevated lead exposure risks in Lexington City (Diversitydatakids.org, 2025; EPA, 2024; Virginia Department of Health, 2024). These factors are undervalued or down-weighted in the COI (Diversitydatakids.org, 2025).

Our results aligned with previous studies and highlight socioeconomic disparity in southeastern U.S. These unequal conditions reflect long-standing stratification processes, specifically the intersection of racial segregation and concentrated poverty, that continue to limit opportunities for young children (Massey & Denton, 1993; Wilson, 1987). However, our driver analysis revealed that these vulnerabilities are not driven by socioeconomic factors alone. Surprisingly, we found that the identified high-vulnerability clusters are predominantly exposure-driven and exhibit a distinct spatial transition toward co-driven and sensitivity and adaptive capacity-driven peripheries, which highlight the importance of integrating environmental management with social support strategies to reduce early childhood health disparities.

For example, in the Southern Great plains, where extreme heat and drought are the primary drivers, policies such as prioritizing shared climate-resilient infrastructure and early warning networks, should be implemented collaboratively across these counties (Centers for Disease & Prevention, 2024; Program, 2023). Alternatively, in Appalachia, high vulnerability is linked to industrial pollution, particularly significantly elevated toxic releases in Kentucky, and a pervasive risk of lead exposure throughout the region (Zhang et al., 2023). Consequently, effective intervention in Appalachia requires stricter enforcement of emission standards, legacy pollution remediation, and comprehensive housing retrofit programs to abate lead hazards.

Importantly, not all exposure-driven counties exhibit high overall vulnerability index values, as exemplified by counties in Iowa and Nebraska. These areas demonstrate that high environmental exposure can be mitigated by lower sensitivity and stronger adaptive capacity levels. In Iowa, high environmental exposure, primarily driven by lead and drought risks identified by the National Risk Index, is effectively mitigated by robust socioeconomic factors, such as high private health insurance coverage and childcare availability (FEMA, 2021). Similarly, in Nebraska, despite facing significant lead exposure and ozone pollution, counties maintain low overall vulnerability. This is largely attributed to protective socioeconomic conditions, including exceptionally low poverty rates and high educational attainment. These examples demonstrate that strong socioeconomic foundations can partially buffer against environmental exposures to safeguard early childhood health. This highlights that strong socioeconomic factors can partially buffer against environmental exposures to protect early childhood health.

Several limitations should be acknowledged. First, the selection of variables was constrained by data availability. Where data for 2023 were unavailable, substitutes from the most recent available years were employed, which may introduce a degree of temporal mismatch. Similarly, due to data accessibility, the analysis was restricted to the county level rather than finer spatial units, limiting the ability to capture within-county heterogeneity. Second, validation relied primarily on child mortality, leaving other health impacts domains such as cognitive development, chronic conditions, and behavioral health underexplored. Finally, while the index was developed using a robust theoretical and empirical framework, it did not incorporate perspectives from stakeholders such as parents, community organizations, or policymakers, which may reduce its immediate applicability in practice.

## 5. Conclusion

This study developed and validated an Early Childhood Environmental Health Vulnerability Index (EC-EHVI) that integrates ESA and Bronfenbrenner’s bioecological framework. By combining theoretical foundations with empirical evidence, the EC-EHVI offers a systematic and comprehensive approach for identifying counties where young children may experience disproportionately cumulative environmental health risks. The results highlight both spatial disparities and key drivers of vulnerability, providing insights into the complex interactions between environmental exposures and social determinants of child health. While the analysis was constrained by data availability and scale, by highlighting spatial inequalities and their underlying drivers, the EC-EHVI offers a valuable decision-support tool to help prioritize resources, regulatory actions, and targeted place-based interventions in high-risk counties. With continued refinement and broader validation, the EC-EHVI can inform targeted interventions, incorporating the EC-EHVI into federal, state, and community planning efforts could help better align environmental, health, and social policies to safeguard the health and well-being of children in vulnerable communities.

## Data Availability

All data produced in the present study are available upon reasonable request to the authors

## Acknowledgments

This work was supported by the EPA (Grant No. R840630). All authors acknowledge the contributions of all participants involved in this research.

## Open Research

The derived data and analysis code generated in this study are publicly available at Zenodo (DOI: 10.5281/zenodo.18751190). All underlying input data are publicly available as described in the Methods section.

## Reference

Achakulwisut, P., Brauer, M., Hystad, P., & Anenberg, S. C. (2019). Global, national, and urban burdens of paediatric asthma incidence attributable to ambient NO(2) pollution: estimates from global datasets. Retrieved from: https://www.ncbi.nlm.nih.gov/pubmed/30981709 https://www.thelancet.com/pdfs/journals/lanplh/PIIS2542-5196(19)30046-4.pdf

U.S. Environmental Protection Agency. (2014). EPA National Walkability Index: Methodology and User Guide [Dataset]. Retrieved from: https://www.epa.gov/smartgrowth/smart-location-mapping

Andersen, R. M. (1995). Revisiting the Behavioral Model and Access to Medical Care: Does It Matter? Journal of Health and Social Behavior, 36(1), 1–10. https://www.jstor.org/stable/2137284

Aradhya, S., Tegunimataka, A., Kravdal, Ø., Martikainen, P., Myrskylä, M., Barclay, K., & Goisis, A. (2023). Maternal age and the risk of low birthweight and pre-term delivery: a pan-Nordic comparison. International Journal of Epidemiology, 52(1), 156–164. 10.1093/ije/dyac211

Banerjee, S., Carlin, B. P., & Gelfand, A. E. (2004). Hierarchical Modeling and Analysis for Spatial Data: Chapman & Hall/CRC.

Besag, J., York, J., & Mollié, A. (1991). Bayesian Image Restoration with Applications in Spatial Statistics (with Discussion). Annals of the Institute of Statistical Mathematics, 43, 1–59.

Bivand, R. S., Pebesma, E., & Gomez-Rubio, V. (2013). Applied Spatial Data Analysis with R. New York: Springer.

Bivand, R. S., & Wong, D. W. (2018). Comparing implementations of global and local indicators of spatial association. TEST, 27(3), 716–748.

Bronfenbrenner, U., & Morris, P. A. (2007). The bioecological model of human development. In R. M. Lerner (Ed.), Handbook of child psychology: Vol. 1. Theoretical models of human development (6th ed., pp. 793–828). Hoboken, NJ: John Wiley & Sons.

Centers for Disease Control and Prevention. (2024). Regional Health Effects: Southern Great Plains. CDC Climate and Health Program. https://www.cdc.gov/climate-health/php/regions/southerngreatplains.html

Centers for Disease Control and Prevention & Agency for Toxic Substances and Disease Registry. (2024). CDC/ATSDR Social Vulnerability Index 2022 Database [Dataset]. Retrieved from: https://www.atsdr.cdc.gov/placeandhealth/svi/data_documentation_download.html

Centers for Disease Control and Prevention & National Center for Health Statistics. (2024). Multiple Cause of Death on CDC WONDER Online Database, 2018–2023 (Provisional) [Dataset]. Retrieved from: http://wonder.cdc.gov/mcd.html

Centers for Disease Control and Prevention. (2024). PLACES: Local Data for Better Health, Technical Documentation. Retrieved from https://www.cdc.gov/places/methodology/index.html

Cohen, J. (2013). Statistical Power Analysis for the Behavioral Sciences. Burlington: Elsevier Science.

Coleman-Jensen, A., Rabbitt, M. P., Gregory, C. A., & Singh, A. (2023). Household Food Security in the United States in 2023. Retrieved from Washington, DC: https://www.ers.usda.gov/publications/pub-details?pubid=109895

County Health Rankings. (2023). County Health Rankings & Roadmaps (CHR) [Dataset]. Retrieved from https://www.countyhealthrankings.org/

Cutler, D. M., & Lleras-Muney, A. (2006). Education and Health: Evaluating Theories and Evidence. Cambridge, MA: National Bureau of Economic Research.

Diversitydatakids.org. (2025). Child Opportunity Index 3.0-2023 database [Dataset]. Retrieved from: https://www.diversitydatakids.org/research-library/child-opportunity-index/child-opportunity-index-30-2023-census-tract-data

Dunnington, D. (2021). ggspatial: Spatial Data Analysis and Visualization with ggplot2 [Software]. Retrieved from https://CRAN.R-project.org/package=ggspatial

European Space Agency. (2018). Sentinel-5 Precursor TROPOMI: Tropospheric Monitoring Instrument [Dataset]. Retrieved from: https://sentinel.esa.int/web/sentinel/missions/sentinel-5p

Evans, J. D. (1996). Straightforward statistics for the behavioral sciences. Pacific Grove: Brooks/Cole Pub. Co.

Federal Emergency Management Agency. (2021). National Risk Index (NRI) [Dataset]. Retrieved from https://hazards.fema.gov/nri/

Federal Emergency Management Agency. (2022). National Risk Index: Technical Documentation. Retrieved from Washington, DC: https://hazards.fema.gov/nri/technical-documentation

Great Britain. Committee on the Medical Effects of Air Pollutants. (1995). Asthma and outdoor air pollution. London: HMSO.

Health Resources Services Administration. (2022). Area Health Resources Files (AHRF) [Dataset]. Retrieved from https://data.hrsa.gov/topics/health-workforce/ahrf

Homeland Infrastructure Foundation-Level Data. (2022). Daycare Center Locations [Dataset]. Retrieved from https://hifld-geoplatform.opendata.arcgis.com/

Intergovernmental Panel on Climate Change (Ed.) (2001). Climate change 2001: Impacts, adaptation, and vulnerability. Contribution of Working Group II to the third assessment report of the Intergovernmental Panel on Climate Change (Third assessment report ed.). Cambridge, UK: Cambridge University Press.

Internal Revenue Service. (2022). Statistics of Income (SOI): Child Tax Credit. Retrieved from https://www.irs.gov/statistics/soi-tax-stats

Jacobs, D. E., Kelly, T., & Sobolewski, J. (2007). Linking public health, housing, and indoor environmental policy: successes and challenges at local and federal agencies in the United States. Environ Health Perspect, 115(6), 976–982. https://www.ncbi.nlm.nih.gov/pubmed/17589610

Jacobson, H., & Wagner, H. (2024). Impact of Outdoor Air Pollution on Child Health and Well-Being. Retrieved from https://www.nemours.org/content/dam/nemours/shared/collateral/policy-briefs/impact-of-outdoor-air-pollution-on-child-health.pdf

James, G., Witten, D., Hastie, T., Tibshirani, R., & SpringerLink. (2021). An Introduction to Statistical Learning : with Applications in R (Version 2nd 2021.). New York, NY: Springer US : Imprint: Springer.

Khoury, M. J., Beaty, T. H., & Cohen, B. H. (1993). Fundamentals of Genetic Epidemiology. New York: Oxford University Press.

Kronick, R. (2013). Health Insurance Coverage and Access to Care. The New England Journal of Medicine, 369(9), 867–869. https://www.nejm.org/doi/full/10.1056/NEJMe1302107

Landrigan, P. J., Fuller, R., Acosta, N. J. R., Adeyi, O., Arnold, R., Basu, N. N., et al. (2018). The Lancet Commission on pollution and health. Lancet, 391(10119), 462–512. https://www.ncbi.nlm.nih.gov/pubmed/29056410

Leventhal, T., & Brooks-Gunn, J. (2000). The neighborhoods they live in: The effects of neighborhood residence on child and adolescent outcomes. Psychological Bulletin, 126(2), 309–337.

Lewis, P. G. T., Chiu, W. A., Nasser, E., Proville, J., Barone, A., Danforth, C., et al. (2023). Characterizing vulnerabilities to climate change across the United States. Environment International, 172, 107772.

Lindgren, F., Rue, H., & Lindström, J. (2011). An explicit link between Gaussian fields and Gaussian Markov random fields: the SPDE approach. Journal of the Royal Statistical Society: Series B, 73(4), 423–498.

Macias-Konstantopoulos, W. L., Collins, K. A., Diaz, R., Duber, H. C., Edwards, C. D., Hsu, A. P., et al. (2023). Race, Healthcare, and Health Disparities: A Critical Review and Recommendations for Advancing Health Equity. West J Emerg Med, 24(5), 906–918. https://www.ncbi.nlm.nih.gov/pubmed/37788031

Mande, M. (2023). Supplemental Nutrition Assistance Program as a Health Intervention. Current Opinion in Pediatrics, 35(1), 98–104. https://journals.lww.com/co-pediatrics/fulltext/2023/02000/supplemental_nutrition_assistance_program_as_a.8.aspx

Markevych, I., Schoierer, J., Hartig, T., Chudnovsky, A., Hystad, P., Dzhambov, A. M., et al. (2017). Exploring pathways linking greenspace to health: Theoretical and methodological guidance. Environ Res, 158, 301–317. https://www.ncbi.nlm.nih.gov/pubmed/28672128

Massey, D. S., & Denton, N. A. (1993). American apartheid : segregation and the making of the underclass. Cambridge, Mass.: Harvard University Press.

Multi-Resolution Land Characteristics Consortium. (2019). National Land Cover Database (NLCD) [Dataset]. Retrieved from https://www.mrlc.gov/

Noelke, C., McArdle, N., DeVoe, B., Leonardos, M., Lu, Y., Ressler, R. W., & Acevedo-Garcia, D. (2025a). Child Opportunity Index 3.0 technical documentation. Retrieved from https://www.diversitydatakids.org/research-library/coi-30-technical-documentation

Noelke, C., McArdle, N., DeVoe, B., Leonardos, M., Lu, Y., Ressler, R. W., & Acevedo-Garcia, D. (2025b). Child Opportunity Index 3.0 Technical Documentation (July 24, 2025). Retrieved from

Pebesma, E. (2018). Simple Features for R: Standardized Support for Spatial Vector Data. The R Journal, 10(1), 439–446.

PolicyMap. (2022). Out-of-Pocket Medical Costs Dataset [Dataset]. Retrieved from: https://www.policymap.com/

U.S. Global Change Research Program. (2023). Fifth National Climate Assessment: Chapter 26 — Southern Great Plains. Retrieved from https://toolkit.climate.gov/sites/default/files/2025-07/NCA5_Ch26_Southern-Great-Plains_Handout.pdf

R Core Team. (2025). R: A language and environment for statistical computing [Software]. Vienna, Austria: R Foundation for Statistical Computing. Retrieved from https://www.R-project.org/

Rappold, A. G., Reyes, J., Pouliot, G., Cascio, W. E., & Diaz-Sanchez, D. (2017). Community Vulnerability to Health Impacts of Wildland Fire Smoke Exposure. Environ Sci Technol, 51(12), 6674–6682. https://www.ncbi.nlm.nih.gov/pubmed/28493694

Ross, C. E., & Wu, C.-l. (1995). The Links Between Education and Health. American Sociological Review, 60(5), 719–745. https://www.jstor.org/stable/2096319

Rue, H., Lindgren, F., & Krainski, E. T. (2022). INLA: Full Bayesian Analysis of Latent Gaussian Models Using Integrated Nested Laplace Approximations [Software]. R Foundation for Statistical Computing. https://www.r-inla.org/

Rue, H., Martino, S., & Chopin, N. (2009). Approximate Bayesian Inference for Latent Gaussian Models Using Integrated Nested Laplace Approximations (with Discussion). Journal of the Royal Statistical Society B, 71, 319–392.

Smit, B., Pilifosova, O., Burton, I., Challenger, B., Huq, S., Klein, R. J. T., et al. (2001). Adaptation to climate change in the context of sustainable development and equity. In J. J. McCarthy, O. F. Canziani, N. A. Leary, D. J. Dokken, & K. S. White (Eds.), Climate change 2001: Impacts, adaptation, and vulnerability. Contribution of Working Group II to the third assessment report of the Intergovernmental Panel on Climate Change (pp. 877–912). Cambridge, UK: Cambridge University Press.

Tee Lewis, P. G., Chiu, W. A., Nasser, E., Proville, J., Barone, A., Danforth, C., et al. (2023). Characterizing vulnerabilities to climate change across the United States. Environ Int, 172, 107772. https://www.ncbi.nlm.nih.gov/pubmed/36731185

The Annie E. Casey Foundation. (2024). 2024 KIDS COUNT data book: 35th Edition. Retrieved from Baltimore, MD: https://www.aecf.org/resources/2024-kids-count-data-book

U.S. Census Bureau. (2022). American Community Survey (ACS) [Dataset]. Retrieved from https://www.census.gov/programs-surveys/acs

U.S. Census Bureau. (2024). American Community Survey Data: Technical Documentation. Retrieved from https://www.census.gov/programs-surveys/acs/technical-documentation.html

U.S. Centers for Disease Control Prevention. (2022). PLACES: Local Data for Better Health (CDC PLACES) [Dataset]. Retrieved from: https://www.cdc.gov/places/

U.S. Department of Agriculture. (2022). Food Security Supplement (FSS) [Dataset]. Retrieved from https://www.ers.usda.gov/topics/food-nutrition-assistance/food-security-in-the-u-s/

U.S. Environmental Protection Agency. (2016). Technical Information about Fused Air Quality Surface Using Downscaling Tool: Metadata description [Dataset]. Retrieved from https://www.epa.gov/sites/default/files/2016-07/documents/data_fusion_meta_file_july_2016.pdf

U.S. Environmental Protection Agency. (2020a). National Emissions Inventory (NEI) [Dataset]. Retrieved from https://www.epa.gov/air-emissions-inventories

U.S. Environmental Protection Agency. (2020b). Smart Location Database (SLD) [Dataset]. Retrieved from https://www.epa.gov/smartgrowth/smart-location-mapping

U.S. Environmental Protection Agency. (2020c). Smart Location Database (SLD): Walkability Index [Dataset]. Retrieved from https://www.epa.gov/smartgrowth/smart-location-mapping

U.S. Environmental Protection Agency. (2022). Safe Drinking Water Information System (SDWIS) [Dataset]. Retrieved from https://www.epa.gov/sdwa

U.S. Environmental Protection Agency. (2023). Federal Air Quality System Data (FAQSD) [Dataset]. Retrieved from https://www.epa.gov/aqs

U.S. Environmental Protection Agency. (2024). Safe Drinking Water Information System (SDWIS) Federal Reporting Services [Dataset]. https://www.epa.gov/ground-water-and-drinking-water/safe-drinking-water-information-system-sdwis-federal-reporting

United Nations International Children’s Emergency Fund. (2025). Children face unique vulnerabilities to environmental hazards at every stage of life. Retrieved from https://www.unicef.org/reports/children-face-unique-vulnerabilities-environmental-hazards-at-every-stage-life

Virginia Department of Health. (2024). Childhood Lead Poisoning Prevention Program (CLPPP): Lead-Safe Virginia. Retrieved from Richmond, VA: https://www.vdh.virginia.gov/leadsafe/

Washington State Department of Health. (2021). Lead Exposure Risk Database [Dataset]. Retrieved from https://doh.wa.gov/

Wickham, H. (2016). ggplot2: Elegant Graphics for Data Analysis. New York: Springer-Verlag.

Williams, D. R., & Collins, C. (2001). Racial residential segregation: A fundamental cause of racial disparities in health. Public Health Reports, 116(5), 404–416.

Wilson, W. J. (1987). The truly disadvantaged : the inner city, the underclass, and public policy (1st paperback ed.). Chicago: University of Chicago Press.

World Health Organization. (2021). WHO global air quality guidelines: particulate matter (PM2.5 and PM10), ozone, nitrogen dioxide, sulfur dioxide and carbon monoxide. Retrieved from Geneva: https://iris.who.int/handle/10665/345329

World Health Organization. (2017). Inheriting a sustainable world: Atlas on children’s health and the environment. Retrieved from Geneva: https://www.who.int/publications/i/item/9789241511773

Zajac, L., Landrigan, P. J., & Council on Environmental Health and Climate Change. (2025). Environmental Issues in Global Pediatric Health: Technical Report. Pediatrics, 155(2). 10.1542/peds.2024-070076

Zhang, C., & Zierold, K. (2023). Exploring Environmental Injustice in Exposure to Airborne Lead from Industrial Facilities in Kentucky. Retrieved from https://www.epa.gov/system/files/documents/2023-11/zhang-charlie_exploring-environmental-injustice-in-exposure-to-airborne-lead-from-industrial-facilities-in-kentucky.pdf

